# Vitamin D insufficiency is prevalent in severe COVID-19

**DOI:** 10.1101/2020.04.24.20075838

**Authors:** Frank H. Lau, Rinku Majumder, Radbeh Torabi, Fouad Saeg, Ryan Hoffman, Jeffrey D. Cirillo, Patrick Greiffenstein

## Abstract

**Background:** COVID-19 is a major pandemic that has killed more than 196,000 people. The COVID-19 disease course is strikingly divergent. Approximately 80-85% of patients experience mild or no symptoms, while the remainder develop severe disease. The mechanisms underlying these divergent outcomes are unclear. Emerging health disparities data regarding African American and homeless populations suggest that vitamin D insufficiency (VDI) may be an underlying driver of COVID-19 severity. To better define the VDI-COVID-19 link, we determined the prevalence of VDI among our COVID-19 intensive care unit (ICU) patients.

**Methods:** In an Institutional Review Board approved study performed at a single, tertiary care academic medical center, the medical records of COVID-19 patients were retrospectively reviewed. Subjects were included for whom serum 25-hydroxycholecalcifoerol (25OHD) levels were determined. COVID-19-relevant data were compiled and analyzed. We determined the frequency of VDI among COVID-19 patients to evaluate the likelihood of a VDI-COVID-19 relationship.

**Results:** Twenty COVID-19 patients with serum 25OHD levels were identified; 65.0% required ICU admission.The VDI prevalence in ICU patients was 84.6%, vs. 57.1% in floor patients. Strikingly, 100% of ICU patients less than 75 years old had VDI. Coagulopathy was present in 62.5% of ICU COVID-19 patients, and 92.3% were lymphocytopenic.

**Conclusions:** VDI is highly prevalent in severe COVID-19 patients. VDI and severe COVID-19 share numerous associations including hypertension, obesity, male sex, advanced age, concentration in northern climates, coagulopathy, and immune dysfunction. Thus, we suggest that prospective, randomized controlled studies of VDI in COVID-19 patients are warranted.

## Introduction

The novel SARS-CoV-2 virus causes COVID-19 and has resulted in 2.8 million confirmed cases and more than 196,000 deaths. Strikingly, 80-85% of patients are asymptomatic or have self-limiting disease.^1^ The remaining require major hospital resources and threaten to collapse our healthcare system. The mechanisms underlying divergent COVID-19 outcomes are unknown.

Emerging health disparities data are potentially illuminating. In Louisiana, African Americans account for 70% of COVID-19 deaths despite representing only 32% of the population.^2^ In a Boston homeless shelter, 100% of 147 COVID-19 positive subjects were asymptomatic.^3^ The mechanisms underlying severe COVID-19 should account for both these findings, as well as other COVID-19 mortality risk factors: hypertension, obesity, male sex, advanced age, concentration in northern climates, and COVID-19 associated coagulopathy (CAC).^4,5^

Vitamin D insufficiency (VDI) meets every one of the above criteria. VDI affects 80-90% of the African American population. In contrast, homeless persons generally have poorer health and nutrition, but can have greater exposure to sunlight, the source of 80-90% of the body’s vitamin D.^6^ VDI causes essential hypertension and is associated with every COVID-19 mortality risk factor.^7,8^ Hydroxychloroquine raises plasma vitamin D levels.^9^ Lastly, VDI induces a prothrombotic state and adversely impacts both innate and adaptive immune responses. To better define the VDI-COVID-19 link, we determined the prevalence of VDI among our COVID-19 intensive care unit (ICU) patients.

## Methods

In an Institutional Review Board approved study performed at a single, tertiary care academic medical center, the medical records of COVID-19 patients between March 27, 2020 and April 21, 2020 were retrospectively reviewed. Subjects were included for whom serum 25-hydroxycholecalcifoerol (25OHD) levels were determined. COVID-19-relevant data were compiled and analyzed. The 25OHD assay was performed in-house, using a UniCel DxI 600 Access Immunoassay System (Beckman Coulter); the laboratory undergoes recertification every six months. VDI was defined as serum 25OHD < 30 ng/mL.^10^

## Results

Twenty COVID-19 patients with serum 25OHD levels were identified; 13 (65.0%) required ICU admission. Overall, few significant differences were identified between ICU and floor patients (Table 1) but statistical analysis was limited by the small number of subjects. Lactate dehydrogenase on admission was significantly higher among ICU patients (441.8 vs. 223.0, p=0.001), consistent with previous reports. No patients were diagnosed with stroke, myocardial infarction, or pulmonary embolus. Two patients (10%) died during the study period.

**Table 1.**
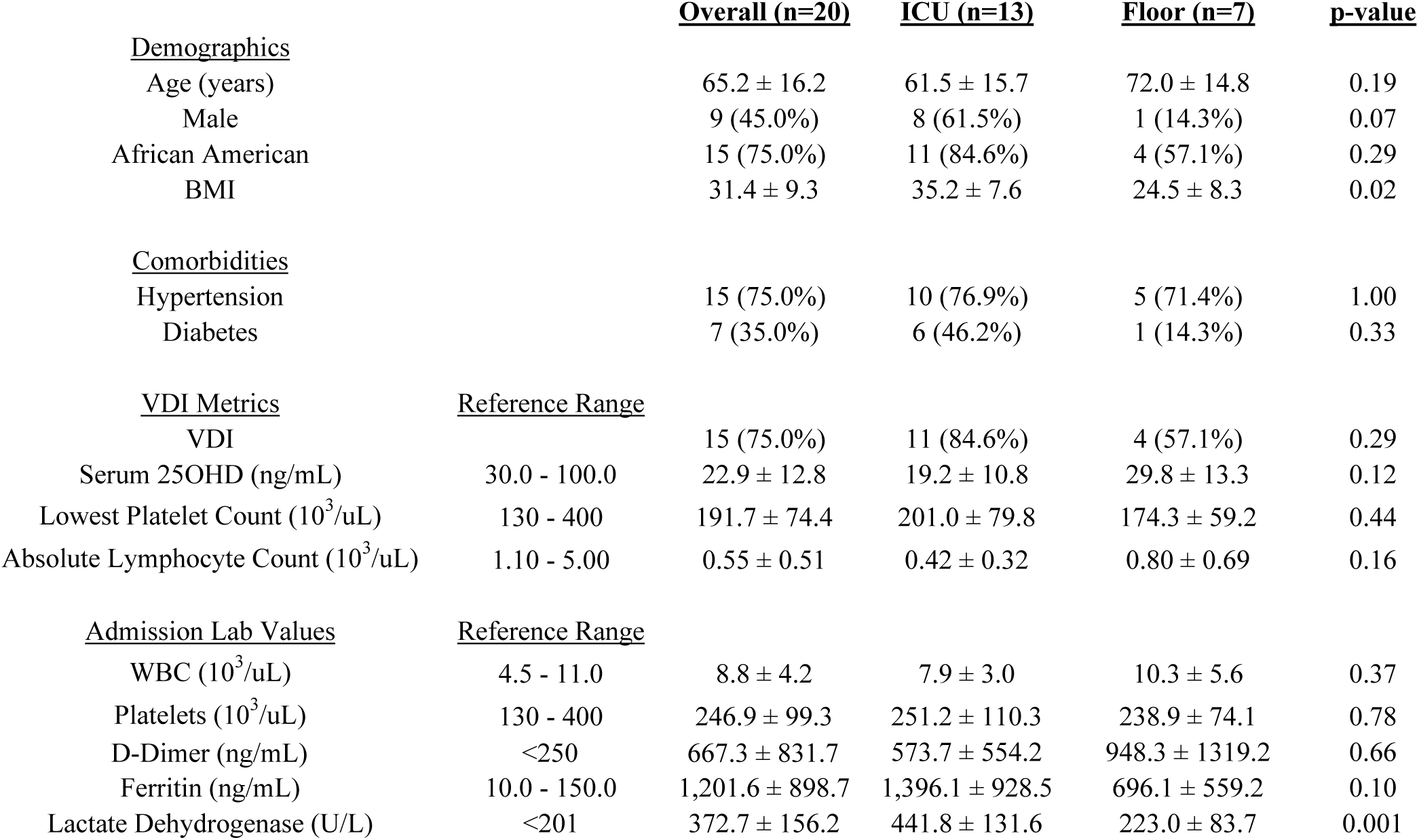
VDI and COVID-19 metrics in ICU vs. floor patients. Values are reported as mean ± standard deviation, or as counts and (%). Tests of significance were Student’s t-test for continuous variables and Fisher’s exact test for categorical variables.

|  |  | **<u>Overall (n=20)</u>** | **<u>ICU (n=13)</u>** | **<u>Floor (n=7)</u>** | **<u>p-value</u>** |
| --- | --- | --- | --- | --- | --- |
| **<u>Demographics</u>** |  |  |  |  |  |
| Age (years) |  | 65.2 ± 16.2 | 61.5 ± 15.7 | 72.0 ± 14.8 | 0.19 |
| Male |  | 9 (45.0%) | 8 (61.5%) | 1 (14.3%) | 0.07 |
| African American |  | 15 (75.0%) | 11 (84.6%) | 4 (57.1%) | 0.29 |
| BMI |  | 31.4 ± 9.3 | 35.2 ± 7.6 | 24.5 ± 8.3 | 0.02 |
| **<u>Comorbidities</u>** |  |  |  |  |  |
| Hypertension |  | 15 (75.0%) | 10 (76.9%) | 5 (71.4%) | 1.00 |
| Diabetes |  | 7 (35.0%) | 6 (46.2%) | 1 (14.3%) | 0.33 |
| **<u>VDI Metrics</u>** |  |  |  |  |  |
|  | **<u>Reference Range</u>** |  |  |  |  |
| VDI |  | 15 (75.0%) | 11 (84.6%) | 4 (57.1%) | 0.29 |
| Serum 25OHD (ng/mL) | 30.0 - 100.0 | 22.9 ± 12.8 | 19.2 ± 10.8 | 29.8 ± 13.3 | 0.12 |
| Lowest Platelet Count (10<sup>3</sup>/uL) | 130 - 400 | 191.7 ± 74.4 | 201.0 ± 79.8 | 174.3 ± 59.2 | 0.44 |
| Absolute Lymphocyte Count (10<sup>3</sup>/uL) | 1.10 - 5.00 | 0.55 ± 0.51 | 0.42 ± 0.32 | 0.80 ± 0.69 | 0.16 |
| **<u>Admission Lab Values</u>** |  |  |  |  |  |
|  | **<u>Reference Range</u>** |  |  |  |  |
| WBC (10<sup>3</sup>/uL) | 4.5 - 11.0 | 8.8 ± 4.2 | 7.9 ± 3.0 | 10.3 ± 5.6 | 0.37 |
| Platelets (10<sup>3</sup>/uL) | 130 - 400 | 246.9 ± 99.3 | 251.2 ± 110.3 | 238.9 ± 74.1 | 0.78 |
| D-Dimer (ng/mL) | <250 | 667.3 ± 831.7 | 573.7 ± 554.2 | 948.3 ± 1319.2 | 0.66 |
| Ferritin (ng/mL) | 10.0 - 150.0 | 1,201.6 ± 898.7 | 1,396.1 ± 928.5 | 696.1 ± 559.2 | 0.10 |
| Lactate Dehydrogenase (U/L) | <201 | 372.7 ± 156.2 | 441.8 ± 131.6 | 223.0 ± 83.7 | 0.001 |

Among ICU subjects, 11 (84.6%) had VDI, vs. 4 (57.1%) of floor subjects. Strikingly, 100% of ICU patients less than 75 years old had VDI (n=11; Table 2). Among these, 64.6% (n=7) had critically low 25OHD (<20 ng/mL) and three had <10 ng/mL. The sepsis-induced coagulopathy score (SIC) was calculable for 8 subjects; 62.5% (n=5) had SIC ≥ 4. Suppressed immune function was prevalent: 92.3% (n=12) were lymphocytopenic, and 9 were profoundly so (absolute lymphocyte count ≤ 0.4 10^3/uL; normal range 1.10-5.00).

**Table 2.**
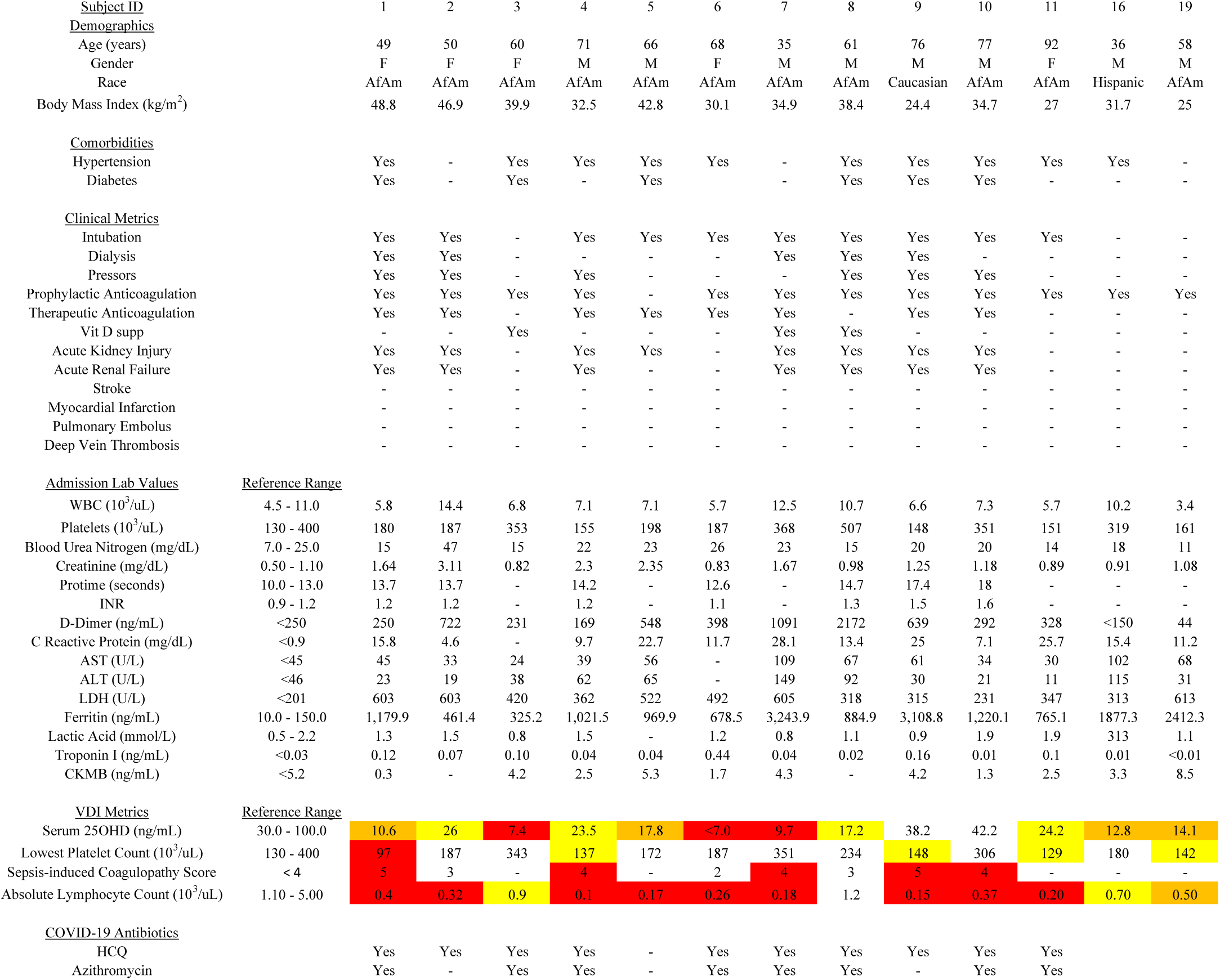
Vitamin D insufficiency, COVID-19-associated coagulopathy, and lymphocytopenia are prevalent in COVID-19 ICU patients. VDI Metrics color coding: yellow = abnormal, orange = highly abnormal, red = critical value. Abbreviations: WBC, white blood count; INR, international normalized ratio; AST, aspartate aminotransferase; ALT, alanine aminotransferase; LDH, lactate dehydrogenase’ CKMB, creatine kinase myocardial band; 25OHD, 25-hydroxycholecalcifoerol; VDI, vitamin D insufficiency.

## Discussion

COVID-19 is an emerging disease whose pathogenic mechanisms are not well understood. Despite being an acute respiratory infection (ARI), its mortality risk factors overlap those of cardiovascular disease: hypertension, diabetes, obesity, advanced age, and male sex. From a health disparities perspective, notable features include an over-representation of African Americans among COVID-19 deaths, and a 100% asymptomatic presentation in a universal survey of a Boston homeless shelter.

Interestingly, VDI and COVID-19 share prevalence patterns for hypertension, diabetes, obesity, advanced age, and male sex (Table 3). VDI can contribute to our understanding of COVID-19 health disparities: VDI is highly prevalent in dark-skinned persons (82.1% of African Americans vs. 41.6% overall). In contrast, although U.S. homeless persons are generally considered to have poor health and decreased access to micronutrients that confer immune benefits, they usually have more exposure to sunlight, a key source of vitamin D production. In Europe, COVID-19 has been severe in Italy, Spain and Greece, but much less so in Scandinavian countries – this precisely recapitulates VDI data showing that Italy, Spain and Greece have VDI rates of 70-90%, vs. 15-30% in Norway and Denmark.^11^ Scandinavian diets contain more vitamin D due to higher fatty fish intake and dairy products supplementation with vitamin D.^11^

**Table 3.** Similarities in demographic and risk factor associations between COVID-19 and Vitamin D Insufficiency. RR, relative risk; 25OHD, 25-hydroxycholecalcifoerol; CI, confidence interval; BMI, body mass index.

| Associations | COVID-19 | Vitamin D Insufficiency |
| --- | --- | --- |
| African American | 70.5% of Louisiana deaths<sup>2</sup> | 82.1% prevalence<sup>8</sup> |
| Elderly (>65 years) | 2.7 - 3.7x hospitalization rates<sup>5</sup> | Prevalence: up to 100%<sup>8</sup> |
| Hypertension | Prevalence: 49.7% of hospitalized patients<sup>5</sup> | Pooled RR of incident hypertension per 10 ng/mL increment in baseline 25OHD levels: 0.88 (95% CI: 0.81-0.97)<sup>7</sup> |
| Obesity | Prevalence: 48.3% of hospitalized patients<sup>5</sup> | Prevalence Ratio (vs. normal BMI): 1.35 (95% CI: 1.21–1.50)<sup>6</sup> |
| Male:Female Ratio | 1.24<sup>5</sup> | 1.44<sup>8</sup> |

The baseline prevalence of VDI amongst ICU patients is 30-40%.^12^ In this study, we found that 84.6% of COVID-19 ICU patients had VDI, vs. 57.1% of floor patients. Strikingly, 100% of ICU patients less than 75 years old had VDI. We also found that 62.5% had CAC, and 92.3% had lymphopenia. Given these data, we hypothesize that VDI enhances COVID-19 severity via 1) its prothrombotic effects and 2) its derangement of the immune response.

### Prothrombosis

CAC is emerging as a key process in severe COVID-19. The American Society of Hematology recommends routine DVT prophylaxis for all admitted COVID-19 patients.^13^ In Wuhan, CAC was present in 71.4% of non-survivors vs. 0.6% in survivors.^14^ Non-survivors demonstrated significantly lower fibrinogen and antithrombin levels on admission, consistent with coagulation factor depletion induced by a hypercoagulable state. A meta-analysis of 1,779 COVID-19 patients reported that platelet counts were significantly lower in severe COVID-19, and that lower platelet counts were associated with mortality.^15^ Anticoagulation can lower mortality: in patients with high SIC scores or D-dimer levels >6-fold the upper limit of normal, heparin reduced mortality to 40.0% vs. 64.2% in controls.^16^ CAC’s role is further evidenced by the multiorgan, microvascular clots in hospitalized COVID-19 patients, which include deep vein thromboses/pulmonary emboli, acute renal failure, cerebrovascular events, myocardial injury, ischemic stroke, and ischemic skin changes.^4^ Microthromboses are found in extrapulmonary organs at a rate greater than in severe acute respiratory syndrome (SARS), another novel coronavirus.^17^

VDI is prothrombotic, since Vitamin D receptor knockout (VDRKO) mice develop a CAC-like response to injury, with aggravated, multiorgan thrombosis following lipopolysaccharide injection.^18^ Expression of antithrombin in the liver and thrombomodulin in the aorta, liver, and kidney were downregulated, whereas tissue factor expression was upregulated in the liver and kidney. In humans, VDI is associated with increased risk of CVD and death.^8^ Vitamin D receptor knockout (VDRKO) mice exhibited increased thrombogenic activity and increased ADP-induced platelet aggregation.

VDR exist in all major cardiovascular cell types, including cardiomyocytes, arterial wall cells, and immune cells. Studies have established that vitamin D metabolites are integral to vascular function and disease, including inflammation and thrombosis. For example, 1,25(OH)2D exerts anticoagulant effects by upregulating the expression of thrombomodulin (an anticoagulant glycoprotein) and downregulating the expression of tissue factor (a critical coagulation factor) in monocytes and human aortic smooth muscle cells.^19^

### Deranged Immunity

Lymphocytopenia is a hallmark of severe COVID-19, suggesting a deranged immune response. Convalescent plasma therapy, a form of passive therapy, can improve the deranged response. Convalescent therapy may improve the host response by reducing VDI or through a protective humoral response. Differentiating these two possibilities is critical to identifying strategies for mitigating severe COVID-19.

VDI leads to deranged immune response, including to viral ARIs. In a study of over 14,000 individuals, VDI was associated with a 58% increase in ARI after controlling for seasonal, demographic, and clinical factors.^20^ A form of macrophage activation syndrome (MAS), where macrophages are highly activated from the initial systemic inflammatory response to SARS-CoV, could be responsible for hyperferritinemia and lead to or exacerbate VDI.^21^ It has been proposed that uncontrolled inflammation commonly leads to hyperferritinemia and likely results in immune dysregulation.^22^ Hyperferritinemia appears to be due to the hemophagocytosis and hypercytokinemia observed in MAS. This clinical picture can result in a prothrombotic state that is consistent with hemophagocytic lymphohistiocytosis and can be observed as a consequence of viral infection.^23^ Overall, these data suggest that an overly exuberant inflammatory response leads to VDI, hyperferritinemia and the prothrombotic state observed with COVID-19.

Vitamin D plays an essential role in modulating both the innate and adaptive immune response.^24,25^ If VDI correlates with severe COVID-19, it would likely explain the high frequency of severe disease in the >60 year old and African American population. Vitamin D-dependent antimicrobial pathways are induced in response to double-stranded RNA, as produced during SARS-CoV-2 replication.^26^ These pathways upregulate antimicrobial peptides, including cathelicidin and β-defensin, and autophagy. In macrophages and endothelial cells, cathelicidin production is modulated in a vitamin D dose-dependent manner.^27–29^ IFN-γ is strongly antimicrobial and a key activator of these pathways, particularly for macrophages and other phagocytic cells, resulting in greater production of reactive oxygen through an oxidative burst and nitrogen species, in addition to these antimicrobial peptides. The over-utilization of these pathways by the host response to SARS-CoV-2 designed to control viral replication could be one mechanism by which VDI arises initially, but once VDI is present, this response becomes ineffective.

VDI prevents the ability of the host to activate these host defensive pathways, but has also been shown to play an important role in macrophage and lymphocyte migration.^30^ Interestingly, the bioactive form of vitamin D, 1,25-dihydroxyvitamin D3 prevents experimental autoimmune encyphalomyelitis (EAE), suggesting that one aspect of the pathology associated with the final stages of deranged immunity observed in COVID-19 may be EAE through inflammation caused by uncontrolled trafficking of macrophages and T cells into the CNS. The massive influx of macrophages and T cells into peripheral organs, including the CNS, may represent the mechanism by which lymphocytopenia arises and could suggest an autoimmune component to the disease. The concept that COVID-19 may impact the CNS is supported by reports of loss of smell and taste by patients and the elevated incidence of ischemic stroke.^31,32^

### The Case Against VDI

While VDI is associated more frequently with ARIs, Vitamin D supplementation does not consistently show benefit against influenza.^33^ However, these outbreaks were not marked by coagulopathy. Furthermore, previous trials did not identify subjects with VDI, thereby introducing a major confounding variable.

## Conclusions

This small, retrospective observational study suggests a link between VDI and severe COVID-19. Anecdotal and observational data indicate that VDI may play a significant role in the progression of the COVID-19 disease state. Low-risk, high-reward potential therapies that target CAC and VDI merit further investigation. Prospective, randomized controlled studies that properly risk-stratify subjects should be performed.

## Data Availability

The authors confirm that the data supporting the findings of this study are available within the article.

## Acknowledgements

We appreciate the University Medical Center of New Orleans for providing access to their patient data.

## References

1. Wu, Z. & McGoogan, J. M. Characteristics of and Important Lessons From the Coronavirus Disease 2019 (COVID-19) Outbreak in China: Summary of a Report of 72 314 Cases From the Chinese Center for Disease Control and Prevention. JAMA (2020) doi:10.1001/jama.2020.2648.

2. Turk, S. Racial disparities in Louisiana’s COVID-19 death rate reflect systemic problems. WWLTV (2020).

3. Baggett, T. P., Keyes, H., Sporn, N. & Gaeta, J. M. COVID-19 outbreak at a large homeless shelter in Boston: Implications for universal testing. medRxiv 2020.04.12.20059618 (2020) doi:10.1101/2020.04.12.20059618.

4. Wadman, M. et al. How does coronavirus kill? Clinicians trace a ferocious rampage through the body, from brain to toes. Science | AAAS https://www.sciencemag.org/news/2020/04/how-does-coronavirus-kill-clinicians-trace-ferocious-rampage-through-body-brain-toes (2020).

5. Garg, S. Hospitalization Rates and Characteristics of Patients Hospitalized with Laboratory-Confirmed Coronavirus Disease 2019 — COVID-NET, 14 States, March 1–30, 2020. MMWR Morb Mortal Wkly Rep 69, (2020).

6. Pereira-Santos, M., Costa, P. R. F., Assis, A. M. O., Santos, C.a.S.T. & Santos, D. B. Obesity and vitamin D deficiency: a systematic review and meta-analysis. Obesity Reviews 16, 341–349 (2015).

7. Kunutsor, S. K., Apekey, T. A. & Steur, M. Vitamin D and risk of future hypertension: meta-analysis of 283,537 participants. Eur. J. Epidemiol. 28, 205–221 (2013).

8. Forrest, K. Y. Z. & Stuhldreher, W. L. Prevalence and correlates of vitamin D deficiency in US adults.Nutrition Research 31, 48–54 (2011).

9. Ruiz-Irastorza, G., Egurbide, M. V., Olivares, N., Martinez-Berriotxoa, A. & Aguirre, C. Vitamin D deficiency in systemic lupus erythematosus: prevalence, predictors and clinical consequences. Rheumatology (Oxford) 47, 920–923 (2008).

10. Holick, M. F. et al. Evaluation, treatment, and prevention of vitamin D deficiency: an Endocrine Society clinical practice guideline. J. Clin. Endocrinol. Metab. 96, 1911–1930 (2011).

11. Scharla, S. H. Prevalence of subclinical vitamin D deficiency in different European countries.Osteoporos Int 8 Suppl 2, S7–12 (1998).

12. Amrein, K., Schnedl, C., Berghold, A., Pieber, T. R. & Dobnig, H. Correction of vitamin D deficiency in critically ill patients - VITdAL@ICU study protocol of a double-blind, placebo-controlled randomized clinical trial. BMC Endocrine Disorders 12, 27 (2012).

13. COVID-19 and Coagulopathy - Hematology.org. https://www.hematology.org:443/covid-19/covid-19-and-coagulopathy.

14. Tang, N., Li, D., Wang, X. & Sun, Z. Abnormal coagulation parameters are associated with poor prognosis in patients with novel coronavirus pneumonia. Journal of Thrombosis and Haemostasis 18, 844–847 (2020).

15. Lippi, G., Plebani, M. & Henry, B. M. Thrombocytopenia is associated with severe coronavirus disease 2019 (COVID-19) infections: A meta-analysis. Clinica Chimica Acta 506, 145–148 (2020).

16. Tang, N. et al. Anticoagulant treatment is associated with decreased mortality in severe coronavirus disease 2019 patients with coagulopathy. J. Thromb. Haemost. (2020) doi:10.1111/jth.14817.

17. Chang, J. C. Acute Respiratory Distress Syndrome as an Organ Phenotype of Vascular Microthrombotic Disease: Based on Hemostatic Theory and Endothelial Molecular Pathogenesis. Clin. Appl. Thromb. Hemost. 25, 1076029619887437 (2019).

18. Aihara, K. et al. Disruption of nuclear vitamin D receptor gene causes enhanced thrombogenicity in mice. J. Biol. Chem. 279, 35798–35802 (2004).

19. Wu-Wong, J. R., Nakane, M. & Ma, J. Vitamin D analogs modulate the expression of plasminogen activator inhibitor-1, thrombospondin-1 and thrombomodulin in human aortic smooth muscle cells. J. Vasc. Res. 44, 11–18 (2007).

20. Monlezun, D. J., Bittner, E. A., Christopher, K. B., Camargo, C. A. & Quraishi, S. A. Vitamin D status and acute respiratory infection: cross sectional results from the United States National Health and Nutrition Examination Survey, 2001-2006. Nutrients 7, 1933–1944 (2015).

21. Crayne, C. B., Albeituni, S., Nichols, K. E. & Cron, R. Q. The Immunology of Macrophage Activation Syndrome. Front. Immunol. 10, (2019).

22. Kernan, K. F. & Carcillo, J. A. Hyperferritinemia and inflammation. Int Immunol 29, 401–409 (2017).

23. Ardalan, M. R., Shoja, M. M., Tubbs, R. S., Esmaili, H. & Keyvani, H. Postrenal Transplant Hemophagocytic Lymphohistiocytosis and Thrombotic Microangiopathy Associated with Parvovirus B19 Infection. American Journal of Transplantation 8, 1340–1344 (2008).

24. Liu, P. T. et al. Toll-like receptor triggering of a vitamin D-mediated human antimicrobial response. Science 311, 1770–1773 (2006).

25. Edfeldt, K. et al. T-cell cytokines differentially control human monocyte antimicrobial responses by regulating vitamin D metabolism. Proc. Natl. Acad. Sci. U.S.A. 107, 22593–22598 (2010).

26. Berry, D. J., Hesketh, K., Power, C. & Hyppönen, E. Vitamin D status has a linear association with seasonal infections and lung function in British adults. Br. J. Nutr. 106, 1433–1440 (2011).

27. Hansdottir, S. et al. Respiratory epithelial cells convert inactive vitamin D to its active form: potential effects on host defense. J. Immunol. 181, 7090–7099 (2008).

28. Wang, T.-T. et al. Cutting edge: 1,25-dihydroxyvitamin D3 is a direct inducer of antimicrobial peptide gene expression. J. Immunol. 173, 2909–2912 (2004).

29. Han, H. et al. Prominent changes in blood coagulation of patients with SARS-CoV-2 infection.Clin. Chem. Lab. Med. (2020) doi:10.1515/cclm-2020-0188.

30. Grishkan, I. V., Fairchild, A. N., Calabresi, P. A. & Gocke, A. R. 1,25-Dihydroxyvitamin D3 selectively and reversibly impairs T helper-cell CNS localization. Proc. Natl. Acad. Sci. U.S.A. 110, 21101–21106 (2013).

31. Eliezer, M. et al. Sudden and Complete Olfactory Loss Function as a Possible Symptom of COVID-19. JAMA Otolaryngol Head Neck Surg (2020) doi:10.1001/jamaoto.2020.0832.

32. Li, Y. et al. Acute Cerebrovascular Disease Following COVID-19: A Single Center, Retrospective, Observational Study. https://papers.ssrn.com/abstract=3550025 (2020) xdoi:10.2139/ssrn.3550025.

33. Gruber-Bzura, B. M. Vitamin D and Influenza—Prevention or Therapy? Int J Mol Sci 19, (2018).

